# SARS-CoV-2 Viral Load is Associated with Increased Disease Severity and Mortality

**DOI:** 10.1101/2020.07.15.20131789

**Authors:** Jesse Fajnzylber, James Regan, Kendyll Coxen, Heather Corry, Colline Wong, Alexandra Rosenthal, Daniel Worrall, Francoise Giguel, Alicja Piechocka-Trocha, Caroline Atyeo, Stephanie Fischinger, Andrew Chan, Keith T. Flaherty, Kathryn Hall, Michael Dougan, Edward T. Ryan, Elizabeth Gillespie, Rida Chishti, Yijia Li, Nikolaus Jilg, Dusan Hanidziar, Rebecca M. Baron, Lindsey Baden, Athe M. Tsibris, Katrina A. Armstrong, Daniel R. Kuritzkes, Galit Alter, Bruce D. Walker, Xu Yu, Jonathan Z. Li, for the Massachusetts Consortium for Pathogen Readiness

**Author notes:** **Corresponding Author:** Jonathan Li, MD, Brigham and Women’s Hospital, 65 Landsdowne Street, Rm 421, Cambridge, MA 02139. Contributed equally to this article (Jesse Fajnzylber, James Regan, Kendyll Coxen, Heather Corry). A list of the Massachusetts Consortium for Pathogen Readiness contributors is provided in the Supplementary Appendix. **Disclosures:** Dr. Li has consulted for Abbvie and Jan Biotech. Dr. Alter is the founder of Seromyx Inc.

## Abstract

The relationship between SARS-CoV-2 viral load and risk of disease progression remains largely undefined in coronavirus disease 2019 (COVID-19). We quantified SARS-CoV-2 viral load from participants with a diverse range of COVID-19 severity, including those requiring hospitalization, outpatients with mild disease, and individuals with resolved infection. SARS-CoV-2 plasma RNA was detected in 27% of hospitalized participants and 13% of outpatients diagnosed with COVID-19. Amongst the participants hospitalized with COVID-19, higher prevalence of detectable SARS-CoV-2 plasma viral load was associated with worse respiratory disease severity, lower absolute lymphocyte counts, and increased markers of inflammation, including C-reactive protein and IL-6. SARS-CoV-2 viral loads, especially plasma viremia, were associated with increased risk of mortality. SARS-CoV-2 viral load may aid in the risk stratification of patients with COVID-19 and its role in disease pathogenesis should be further explored.

## INTRODUCTION

In coronavirus disease 2019 (COVID-19), the relationship between levels of viral replication and disease severity remains unclear. In prior analyses of the SARS-CoV-1 outbreak, viral load within the nasopharynx was associated with worsened disease severity and increased mortality^1,2^. However, it is also clear that there are significant differences between SARS-CoV-1 and 2, including differences in the temporal nature of viral shedding^3,4^, transmissibility^5^, epidemiology^6^, and clinical manifestations^7,8^. Additional studies are needed to determine whether the degree of SARS-CoV-2 viral load within the respiratory tract or other compartments may predict disease outcomes.

The need for additional SARS-CoV-2 viral studies is not only limited to respiratory specimens but extends to the blood. Respiratory failure is the primary cause of death in patients with COVID-19, but complications arising from a hyperactive immune response and vascular damage are also prominently featured both in the pulmonary and extrapulmonary systems^9-13^. In addition, there is a suggestion that detectable plasma viremia using a qualitative qPCR assay may correlate with disease severity^14^, although studies to date have been hampered by the lack of viral load quantification. Together, these findings suggest the importance of systemic SARS-CoV-2 viral circulation, but little is known about the prevalence and magnitude of plasma viremia in predicting COVID-19 outcomes. In this study, we quantified SARS-CoV-2 viral load from the respiratory tract, plasma and urine of participants with a diverse range of COVID-19 severity, including individuals requiring hospitalization, symptomatic non-hospitalized participants, and those recovered from COVID-19 disease.

## RESULTS

### Participant characteristics and SARS-CoV-2 viral loads

We enrolled 88 hospitalized participants with COVID-19, 94 symptomatic individuals who were evaluated in a respiratory infection clinic, of whom 16 were diagnosed with COVID-19 by standard clinical testing of nasopharyngeal swabs, and 53 participants diagnosed with COVID-19 who had symptomatically recovered. Table 1 shows baseline demographic information, disease severity and hospital outcomes. Hospitalized participants were significantly older than both symptomatic outpatients and individuals recovered from COVID-19 (Kruskal-Wallis *P*<0.001). Participants recruited in the outpatient setting had the shortest time between the start of symptoms and the time of sample collection (median 5 days) compared to hospitalized individuals (median 13 days) and recovered participants (median 27 days).

**Table 1.** Demographics and Clinical Characteristics of Participants at Baseline.

| Characteristic | Hospitalized<br>(N=88) | Symptomatic Non-Hospitalized (N=90) | Recovered<br>(N=53) |
| --- | --- | --- | --- |
| Female sex, % | 38% | 62% | 65% |
| Age, median years [Q1,Q3] | 57 [43,68] | 48 [31,59] | 33 [29,42] |
| Ethnicity |  |  |  |
| Caucasian | 35% | 79% | 81% |
| Black/African American | 15% | 8% | 6% |
| Hispanic/Latino | 38% | 3% | 6% |
| Other | 12% | 10% | 8% |
| Comorbidities |  |  |  |
| Hypertension | 53% | 22% | 2% |
| Chronic Lung Disease | 18% | 30% | 2% |
| Diabetes | 40% | 11% | 2% |
| BMI |  |  |  |
| <25 | 20% | 35% | 62% |
| 25-29.99 | 35% | 24% | 24% |
| ≥30 | 45% | 40% | 14% |
| Days Between Symptom Onset and Initial Sample<br>Collection, median [Q1,Q3] | 13 [10,18] | 5 [2,15] | 27 [20, 34] |
| Oxygenation Status at Time of Enrollment |  |  |  |
| Room Air | 15% |  |  |
| Nasal Cannula | 37% |  |  |
| Ventilator | 48% |  |  |
| Hospitalization Status |  |  |  |
| % discharged | 85% |  |  |
| % mortality | 13% |  |  |

We report SARS-CoV-2 viral load analysis both as a continuous variable and analyzed as a categorical variable (detectable versus undetectable) given that only qualitative commercial qPCR testing is available for clinical care. Amongst hospitalized individuals, the majority still had detectable SARS-CoV-2 RNA at the time of initial sample collection, including 50% with detectable SARS-CoV-2 RNA by nasopharyngeal swab, 67% by oropharyngeal swab, and 85% by sputum testing. We also performed SARS-CoV-2 viral load testing from specimens outside of the respiratory tract and found that 27% of participants had detectable SARS-CoV-2 plasma viremia and 10% had detectable viral RNA in the urine (Fig 1). In those with detectable plasma viremia, the median viral load was 2.4 log_10_ RNA copies/mL (range 1.8 – 3.8 log_10_ RNA copies/mL), which was significantly lower than that detected in sputum (median 4.4 log_10_ RNA copies/mL, range 1.8 – 9.0 log_10_ RNA copies/mL, Wilcoxon signed-rank *P* < 0.001).

**Figure 1.**
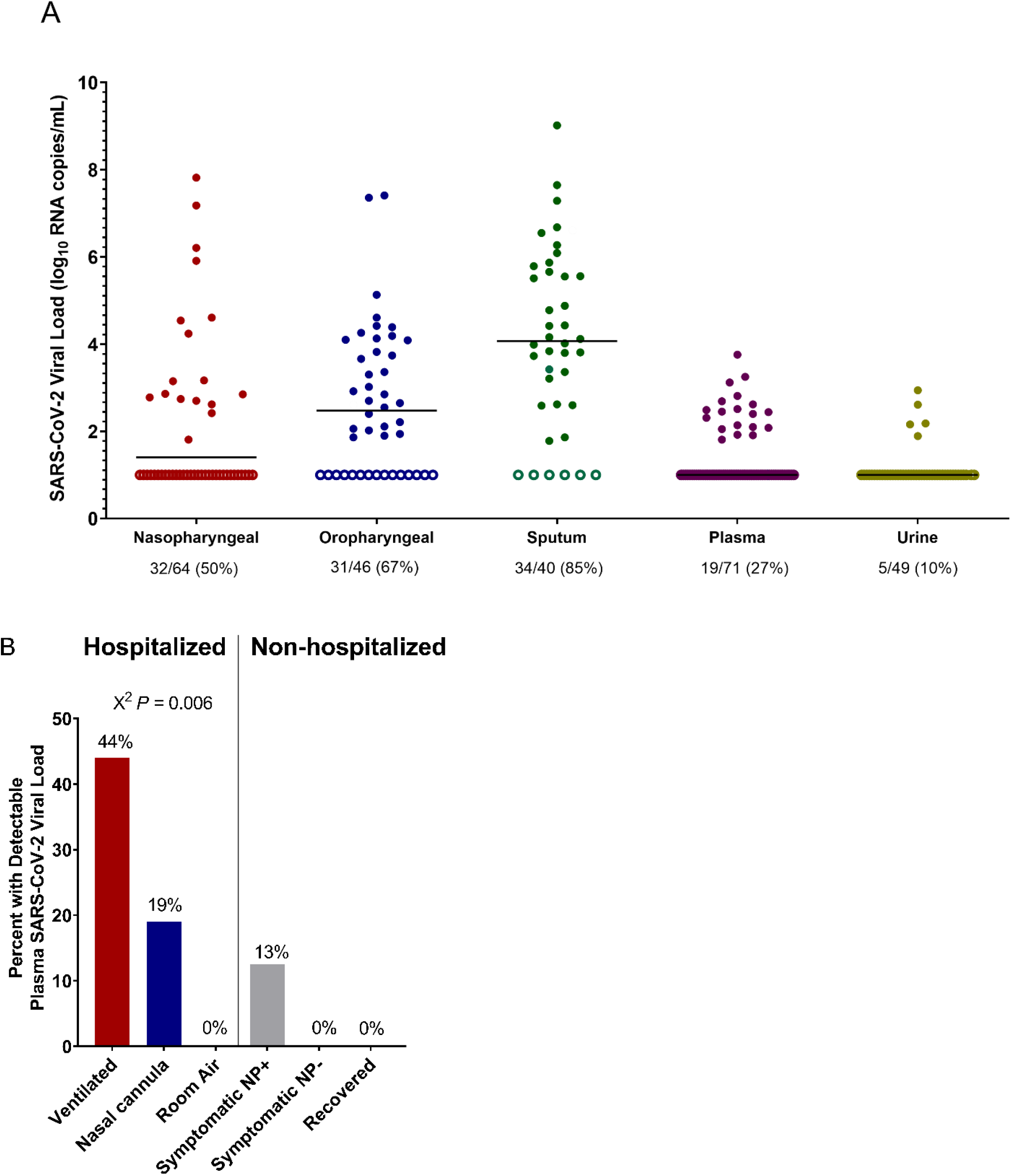
SARS-CoV-2 viral loads at the time of initial sampling. (A) Levels of SARS-CoV-2 viral loads at the time of initial sampling and across specimen types. The percent of samples with detectable viral loads are shown at the bottom. (B) (A) Percent of participants with detectable plasma SARS-CoV-2 viral load by hospitalization status and disease severity. Symptomatic nasopharyngeal swab positive (NP+) and negative (NP-) individuals were evaluated at an outpatient respiratory infection clinic. Recovered individuals included participants who had previously been diagnosed with COVID-19, but whose symptoms have since resolved.

Levels of SARS-CoV-2 viral load were significantly correlated between each of the different respiratory specimen types (nasopharyngeal vs oropharyngeal Spearman *r* = 0.34, *P* = 0.03; nasopharyngeal vs sputum *r* = 0.39, P = 0.03, oropharyngeal vs sputum *r* = 0.56, *P* = 0.001, Supplemental Fig 1). Plasma viral load was modestly associated with both nasopharyngeal (*r* = 0.32, *P* = 0.02) and sputum viral loads (*r* = 0.36, *P* = 0.049), but not significantly associated with oropharyngeal viral loads. There was no significant association between urine viral load and viral loads from any other sample types.

### SARS-CoV-2 viral load is associated with disease severity and laboratory abnormalities

Detectable plasma viremia was generally associated with increased disease severity amongst hospitalized participants as 44% of those on a ventilator had detectable viremia compared to 19% of those receiving supplemental oxygen by nasal cannula and 0% of individuals not requiring supplemental oxygen (X^2^ *P* = 0.006, Fig 1b). Two of the 16 (13%) COVID-19 diagnosed outpatients were found to also have detectable SARS-CoV-2 plasma viremia, compared to none of the 74 outpatients with negative clinical nasopharyngeal testing for SARS-CoV-2 RNA and none of the 53 recovered individuals who had previously been diagnosed with COVID-19. None of the 18 plasma samples from intensive care unit participants collected in the pre-COVID era were found to have detectable plasma SARS-CoV-2 RNA.

In hospitalized participants, higher plasma viral loads were significantly associated with several markers of inflammation and disease severity, including lower absolute lymphocyte counts (Spearman *r* = −0.31, *P* = 0.008), and higher levels of both CRP (*r* = 0.40, *P* < 0.001) and IL-6 (*r* = 0.50, *P* < 0.001). Significant associations were also detected between nasopharyngeal and sputum viral loads and these three markers (Fig 2a). When analyzed as a categorical variable, individuals with detectable plasma, nasopharyngeal or sputum viral loads had significantly lower absolute lymphocyte counts, and higher CRP and IL-6 levels compared to those without detectable plasma viremia (Fig 2b-d). Plasma, nasopharyngeal and/or oropharyngeal viral loads were also significantly associated with increased levels of the inflammatory cytokines IL-8, IP-10, MCP1, IFN-γ, and IL-1RA (Fig 2a).

**Figure 2.**
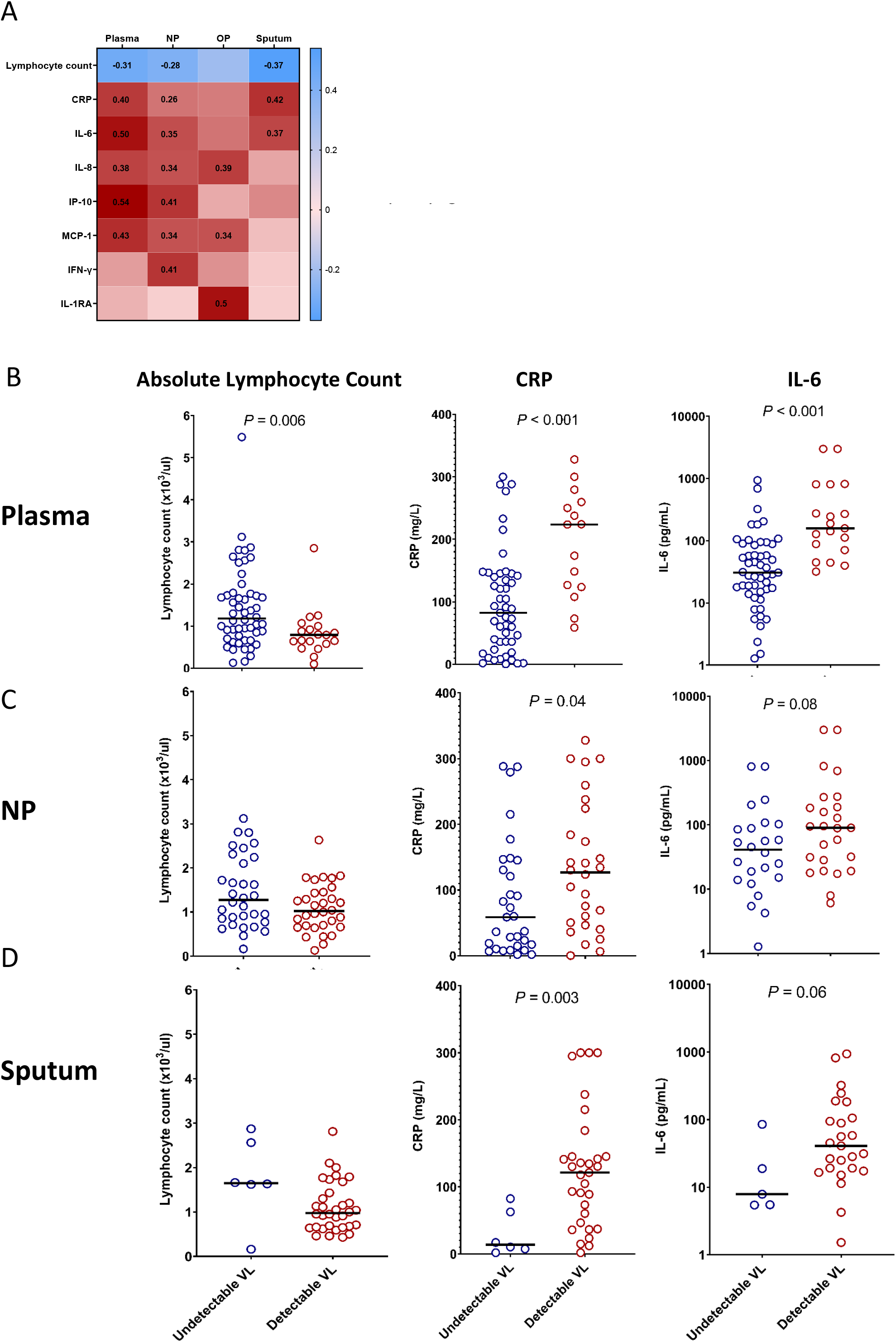
SARS-CoV-2 viral load is associated with markers of inflammation and disease severity. (A) Heat map of spearman correlation values with bold numbers indicating *P* < 0.05. Absolute lymphocyte count, CRP, and IL-6 levels in hospitalized participants with and without detectable plasma (B), nasopharyngeal (C), and sputum (D) viral loads. NP, nasopharyngeal; VL, viral load.

### SARS-CoV-2 viral loads and mortality risk

Compared to individuals who were discharged from the hospital, those who eventually died had significantly higher levels of plasma viremia at the time of initial sampling (median plasma viral load 1.0 vs 2.0 log_10_ RNA copies/mL, *P* = 0.009, Fig 3a), which occurred a median 11 days before death. For hospitalized individuals with initial detectable viremia, 32% died vs 8% of those without initial viremia (OR 5.5, *P* = 0.02, Fig 3e). We performed a sensitivity analysis to assess whether plasma viremia may also predict mortality in those with the most severe disease. For participants who were on ventilatory support at the time of initial sample collection, 43% of those with detectable plasma viremia died compared to 17% of those without detectable plasma viremia, although this comparison did not reach statistical significance (OR 3.8, *P* = 0.11). We also performed an analysis in older participants as the majority of participants who died were at least 70 years old. In those ≥70 years old with initial plasma viremia, 6 of 7 died (86%) vs 2 of 9 (22%) without initial viremia (OR 21, *P* = 0.02). Levels of SARS-CoV-2 viral load in respiratory secretions were also higher in those who eventually died (Fig 3b-d), although the presence or absence of detectable respiratory secretion viral RNA were not significantly associated with increased risk of death (Fig 3f-h). Logistic regression analysis was also performed with viral loads as a continuous variable and plasma, oropharyngeal and sputum viral loads were all associated with increased risk of death (Supplementary Table 1).

**Figure 3.**
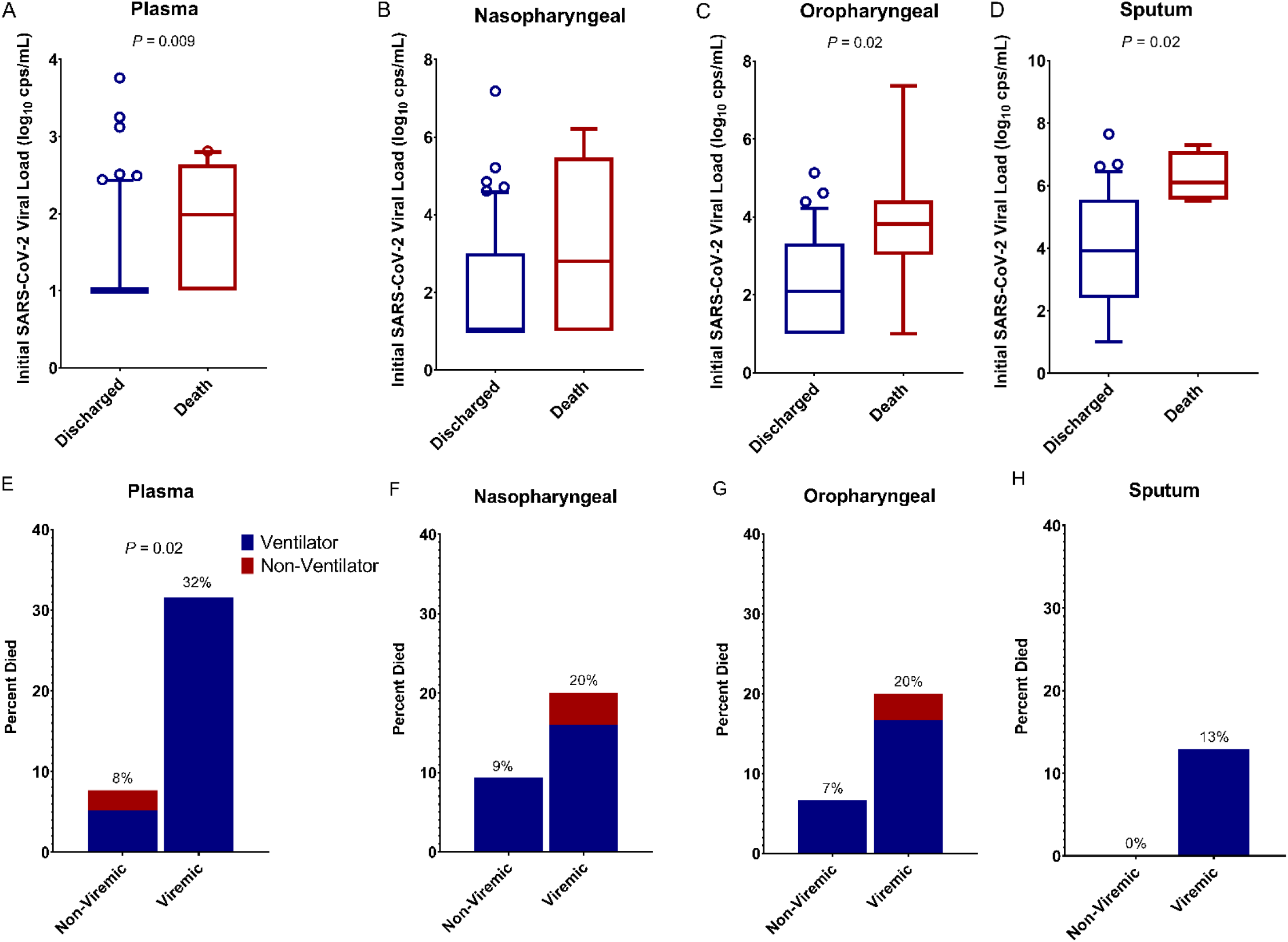
SARS-CoV-2 viral load and risk of death. (A-D) Participants who died had higher initial viral loads compared to those who survived to discharge. (E-H) Percent of participants who eventually died categorized by detectable viral load and disease severity at the time of initial sampling.

A subset of hospitalized participants had longitudinal viral load measurements. Levels of plasma and respiratory viral loads declined from the first and second sampling time points in almost all participants, regardless of eventual participant outcome (Fig 4).

**Figure 4.**
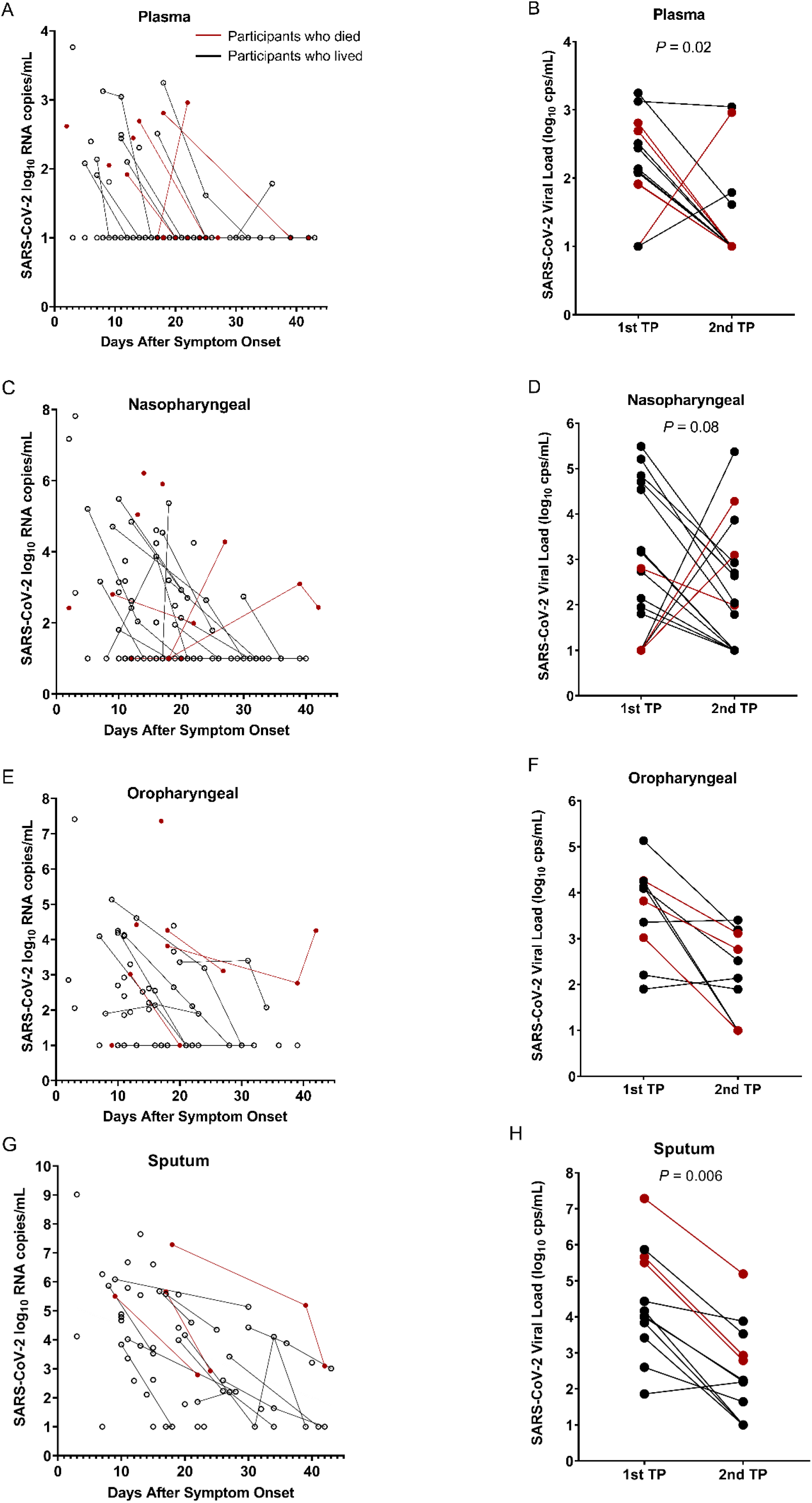
Longitudinal viral load measurements of samples obtained from plasma (A-B), nasopharyngeal swab (C-D), oropharyngeal swab (E-F), or Sputum (G-H). Red dots and lines show viral loads in those who died. Sign test p-values showing significant changes over time are reported in an analysis of viral loads at the first and second available time points (TP).

## DISCUSSION

We report a comprehensive analysis of SARS-CoV-2 respiratory tract, plasma, and urine viral loads of 235 participants who were either hospitalized with COVID-19, evaluated as symptomatic outpatients, or had recovered from COVID-19 disease. The results show a relatively high prevalence of SARS-CoV-2 plasma viremia in hospitalized individuals with severe disease, but plasma viremia was also detected in symptomatic non-hospitalized participants. Levels of SARS-CoV-2 viremia was also associated with markers of inflammation and disease severity, including low lymphocyte counts, and elevated CRP and IL-6 levels. To our knowledge, this is also the first report that SARS-CoV-2 viral loads, especially detectable plasma viremia, predicted the risk of death.

In contrast to prior reports suggesting that the SARS-CoV-2 viral infection is largely confined to the respiratory and gastrointestinal tracts^15,16^, we were able to detect plasma viremia in a substantial proportion of both hospitalized and non-hospitalized participants. The prevalence of SARS-CoV-2 plasma viremia was lower than that found in respiratory secretions, but detectable plasma viremia had a clear relationship with concurrent clinical disease severity, lower absolute lymphocyte count, higher levels of inflammation and increased risk of death. Across the spectrum of viral infections, the extent of viral load has been a predictor of disease severity and progression, including for HIV^17,18^, Ebola^19^, influenza and other non-COVID-19 respiratory viral infections^20-22^. The detection of plasma viral load has been described for both SARS-CoV-1^23,24^ and SARS-CoV-2^14^, but its role in pathogenesis and ability to predict clinical outcomes remains unresolved. To our knowledge, this is the first report demonstrating that SARS-CoV-2 is frequently detectable in plasma and that detectable viral load, both in plasma and the respiratory tract, are associated with increased disease severity and mortality. Therefore, the detection and quantification of viral RNA levels may aid in the risk stratification of patients hospitalized with COVID-19. The association between SARS-CoV-2 viral load with levels of CRP and IL-6 results also suggest that active viral infection could contribute to the hyperinflammatory state that is a hallmark of severe COVID-19^25^. However, the causes of inflammation in COVID-19 could be multifactorial, especially as a subset of participants had elevated inflammatory markers without detectable plasma viremia. Additional studies are needed to determine whether antiviral treatment may effectively interrupt this pathway and whether levels of SARS-CoV-2 viral load could stratify patients into individuals who are more likely to benefit from an antiviral agent versus those with isolated immune dysregulation who may benefit more from an anti-inflammatory or immune-modifying agent^13^.

The source for the plasma viremia is still not fully defined and could reflect spillage from the pulmonary tissue into the vasculature, but there is evidence that SARS-CoV-2 can also directly infect endothelial cells. Angiotensin-converting enzyme 2 (ACE2) is the primary receptor for SARS-CoV-2 and can be found on both arterial and venous endothelial cells^26^ and other perivascular cells^27^. Tissue studies have also revealed evidence of endothelitis with perivascular inflammation^12^ and the extrapulmonary spread of SARS-CoV-2 to other organs^28^. While additional infectivity studies are needed to confirm that plasma viremia represents infectious virions, these previously published support the concept that COVID-19 should be considered more than an isolated respiratory tract infection and that endothelial infection and systemic circulation of infectious SARS-CoV-2 virions may be contributing to the increasing reports of extrapulmonary and micro- and macrovascular complications of COVID-19 that are often disproportionate to the degree of disease severity^9-12,29-32^.

There is an intense search for biomarkers of COVID-19 disease progression that could accelerate early-phase clinical studies of antiviral agents against SARS-CoV-2. There has been an expectation that respiratory tract viral shedding could serve as such a surrogate biomarker, but it is unclear if such assumptions are accurate. An example is the reported clinical benefit of remdesivir^33^ despite the lack of evidence that remdesivir significantly reduces respiratory tract viral loads^34^. We found only modest correlations between respiratory tract viral loads and those of the plasma. This highlights the need for additional studies to assess whether these anatomic compartments may serve as distinct sites of viral replication and whether antiviral medications might have differential effects on viral respiratory tract shedding versus plasma viremia.

Our study has a few notable limitations. First, sputum samples were obtained for only a subset of participants as many participants were unable to generate a sample. While sputum samples had the highest frequency of SARS-CoV-2 detection, this finding demonstrates a potential limitation in their use as a reliable diagnostic modality. Our longitudinal analysis of viral load changes was limited to a subset of participants due to limits on the frequency of blood draws for hospitalized individuals and early discharges in those with relatively mild disease. Additional studies of plasma viral load dynamics early in the course of disease are needed.

In summary, we report that SARS-CoV-2 plasma viremia is commonly detected in hospitalized individuals but can also be detected in symptomatic non-hospitalized outpatients diagnosed with COVID-19. SARS-CoV-2 viral loads, especially within plasma, are associated with systemic inflammation, disease progression, and increased risk of death. The role of SARS-CoV-2 as a mediator of vascular and extrapulmonary COVID-19 disease manifestations should be further explored.

## Data Availability

Data requests can be made to the corresponding author.

## Acknowledgements

We would like to thank the participants of this trial and the dedicated clinical staff who cared for them. We appreciate the support of the Ragon Institute of MGH, MIT and Harvard, the Massachusetts General Hospital Translational and Clinical Research Center, the Brigham and Women’s Hospital Center for Clinical Investigation and the Partners Biobank.

## Appendix

Massachusetts Consortium for Pathogen Readiness (MassCPR) contributors: Betelihem A. (Betty) Abayneh^3^, Patrick Allen^3^, Diane Antille^3^, Alejandro Balazs^2,3^, Julia Bals^2,3^, Max Barbash^2,3^, Yannic Bartsch^2,3^, Julie Boucau^2,3^, Siobhan Boyce^3^, Joan Braley^3^, Karen Branch^3^, Katherine Broderick^3^, Julia Carney^3^, Josh Chevalier^2,3^, Manish C Choudhary^1^, Navin Chowdhury^2,3^, Trevor Cordwell^1^, George Daley^4^, Susan Davidson^3^, Michael Desjardins^1^, Lauren Donahue^1^, David Drew^3^, Kevin Einkauf^2,3^, Sampson Elizabeth^1^, Ashley Elliman^3^, Behzad Etemad^1^, Jon Fallon^2,3^, Liz Fedirko^2,3^, Kelsey Finn^2,3^, Jeanne Flannery^3^, Pamela Forde^3^, Pilar Garcia-Broncano^2,3^, Elise Gettings^3^, David Golan^4^, Kirsten Goodman^1^, Amanda Griffin^3^, Sheila Grimmel^3^, Kathleen Grinke^3^, Ciputra Adijaya Hartana^2,3^, Meg Healy^3^, Howard Heller^3,4^, Deborah Henault^3^, Grace Holland^3^, Chenyang Jiang^2,3^, Hannah Jordan^1^, Paulina Kaplonek^2,3^, Elizabeth W. Karlson^1^, Marshall Karpell^2,3^, Chantal Kayitesi^3^, Evan C. Lam^2,3^, Vlasta LaValle^3^, Kristina Lefteri^2,3^, Xiaodong Lian^2,3^, Mathias Lichterfeld^1,3^, Daniel Lingwood^2,3^, Hang Liu^2,3^, Jinqing Liu^2,3^, Kell Lopez^1^, Yuting Lu^3^, Sarah Luthern^3^, Ngoc L. Ly^2,3^, Maureen MacGowan^1^, Karen Magispoc^1^, Jordan Marchewka^3^, Brittani Martino^3^, Roseann McNamara^3^, Ashlin Michell^2,3^, Ilan Millstrom^2,3^, Noah Miranda^2,3^, Christian Nambu^3^, Susan Nelson^3^, Marjorie Noone^3^, Lewis Novack^1^, Claire O’Callaghan^2,3^, Christine Ommerborn^3^, Matthew Osborn^2,3^, Lois Chris Pacheco^3^, Nicole Phan^3^, Shiv Pillai^2,3^, Falisha A. Porto^3^, Yelizaveta Rassadkina^2,3^, Alexandra Reissis^2,3^, Francis Ruzicka^2,3^, Kyra Seiger^2,3^, Kathleen Selleck^3^, Libera Sessa^2,3^, Arlene Sharpe^4^, Christianne Sharr^2,3^, Sally Shin^2,3^, Nishant Singh^2,3^, Sue Slaughenhaupt^3^, Kimberly Smith Sheppard^3^, Weiwei Sun^2,3^, Xiaoming Sun^2,3^, Elizabeth (Lizzie) Suschana^3^, Opeyemi Talabi^1^, Hannah Ticheli^2,3^, Scott T. Weiss^1^, Vivine Wilson^3^, Alex Zhu^2,3^

1. Brigham and Women’s Hospital, Harvard Medical School, Boston, MA
2. Ragon Institute of MGH, MIT and Harvard, Harvard Medical School, Cambridge, MA
3. Massachusetts General Hospital, Harvard Medical School, Boston, MA
4. Harvard Medical School, Boston, MA

### METHODS

#### Participant enrollment and sample collection

We enrolled hospitalized and non-hospitalized participants with COVID-19 in a longitudinal sample collection study at two academic medical centers. Blood was collected from consented hospitalized participants diagnosed with COVID-19, non-hospitalized symptomatic individuals seeking care at a respiratory infection clinic, and participants who had recovered from known COVID-19 disease. Nasopharyngeal swabs, oropharyngeal swabs, sputum, and urine were also collected from hospitalized participants. Nasopharyngeal swabs and oropharyngeal swabs were collected in 3 mL of phosphate buffered saline (PBS). A subset of hospitalized participants had longitudinal samples collected. Plasma obtained from a cohort of individuals in the intensive care unit from the pre-COVID-19 era were used as a comparator group^35^. Each participant’s electronic medical record was reviewed to determine the oxygenation status (room air, on oxygen by nasal cannula, or requiring ventilator support), demographics, comorbidities and the outcome of the hospitalization (discharge or death). This study was approved by the Partners Institutional Review Board.

#### Markers of inflammation and disease severity

Levels of C-reactive protein (CRP) and absolute lymphocyte count were recorded from the electronic medical record. Thirty-five additional markers of inflammation were evaluated in plasma by the Luminex xMAP assay (ThermoFisher): EGF, Eotaxin, FGF-basic, G-CSF, GM-CSF, HGF, IFN-α, IFN-γ, IL-1α, IL-1β, IL-1RA, IL-2, IL-2R, IL-3, IL-4, IL-5, IL-6, IL-7, IL-8, IL-9, IL-10, IL-12 (p40/p70) IL-13, IL-15, IL-17A, IL-17F, IL-22, IP-10, MCP-1, MIG, MIP-1α, MIP-1β, RANTES, TNF-α, and VEGF.

#### SARS-CoV-2 Viral Load Quantification

Levels of SARS-CoV-2 viral load were quantified using the US CDC 2019-nCoV_N1 primers and probe set^36^. Virions were pelleted from respiratory secretions, swab fluids, plasma, or urine by centrifugation at approximately 21,000 × g for 2 hours at 4^°^C. The supernatant was removed and 750 µL of TRIzol-LS™ Reagent (ThermoFisher) was added to the pellets and then incubated on ice. Following incubation, 200 µL of chloroform (MilliporeSigma) was added and vortexed. The mixtures were separated by centrifugation at 21,000 × g for 15 minutes at 4^°^C, and subsequently the aqueous layer was removed and treated with an equal volume of isopropanol (Sigma). GlycoBlue™ Coprecipitant (ThermoFisher) and 100 µL 3M Sodium Acetate (Life Technologies) were added to each sample and incubated on dry ice until frozen. RNA was pelleted by centrifugation at 21,000 × g for 45 minutes at 4^°^C. The supernatant was discarded and the RNA was washed with cold 70% ethanol. The RNA was resuspended in DEPC-treated water (ThermoFisher).

Each reaction contained extracted RNA, 1X TaqPath™ 1-Step RT-qPCR Master Mix, CG (ThermoFisher), the CDC N1 forward and reverse primers, and probe^36^. Viral copy numbers were quantified using N1 qPCR standards in 16-fold dilutions to generate a standard curve. The assay was run in triplicate for each sample and two non-template control (NTC) wells were included as negative controls. Quantification of the Importin-8 (IPO8) housekeeping gene RNA level was performed to determine the quality of respiratory sample collection. An internal virion control (RCAS) was spiked into each sample and quantified to determine the efficiency of RNA extraction and qPCR amplification^37^.

#### Statistical analyses

Levels of SARS-CoV-2 viral load at the time of initial hospital collection were compared by site of sampling, disease severity and hospital outcome. SARS-CoV-2 viral load analysis was performed both as continuous variables with non-parametric rank-based testing and as a categorical variable (detectable vs undetectable) with Fisher’s exact and X^2^ tests given the qualitative nature of current commercial qPCR tests. SARS-CoV-2 viral loads below 40 RNA copies/mL were categorized as undetectable and set at 1.0 log_10_ RNA copies/mL. For the subset of participants with repeated sampling, the sign test was used to assess viral load change between the first and second time point. Correlation analysis was performed using Spearman rank-based testing. In the correlation analysis between the soluble inflammatory markers and viral load, a P-value <0.01 between a marker and any of the viral load measurements was the threshold to include that marker in the reported results. Logistic regression and other statistical analyses were performed using GraphPad Prism 8 and SAS software, version 9.4. Only univariate analysis was performed due to the available sample size, but we did perform sensitivity analysis for plasma viral load effects based on disease severity and age.

---

**Supplemental Figure 1.**
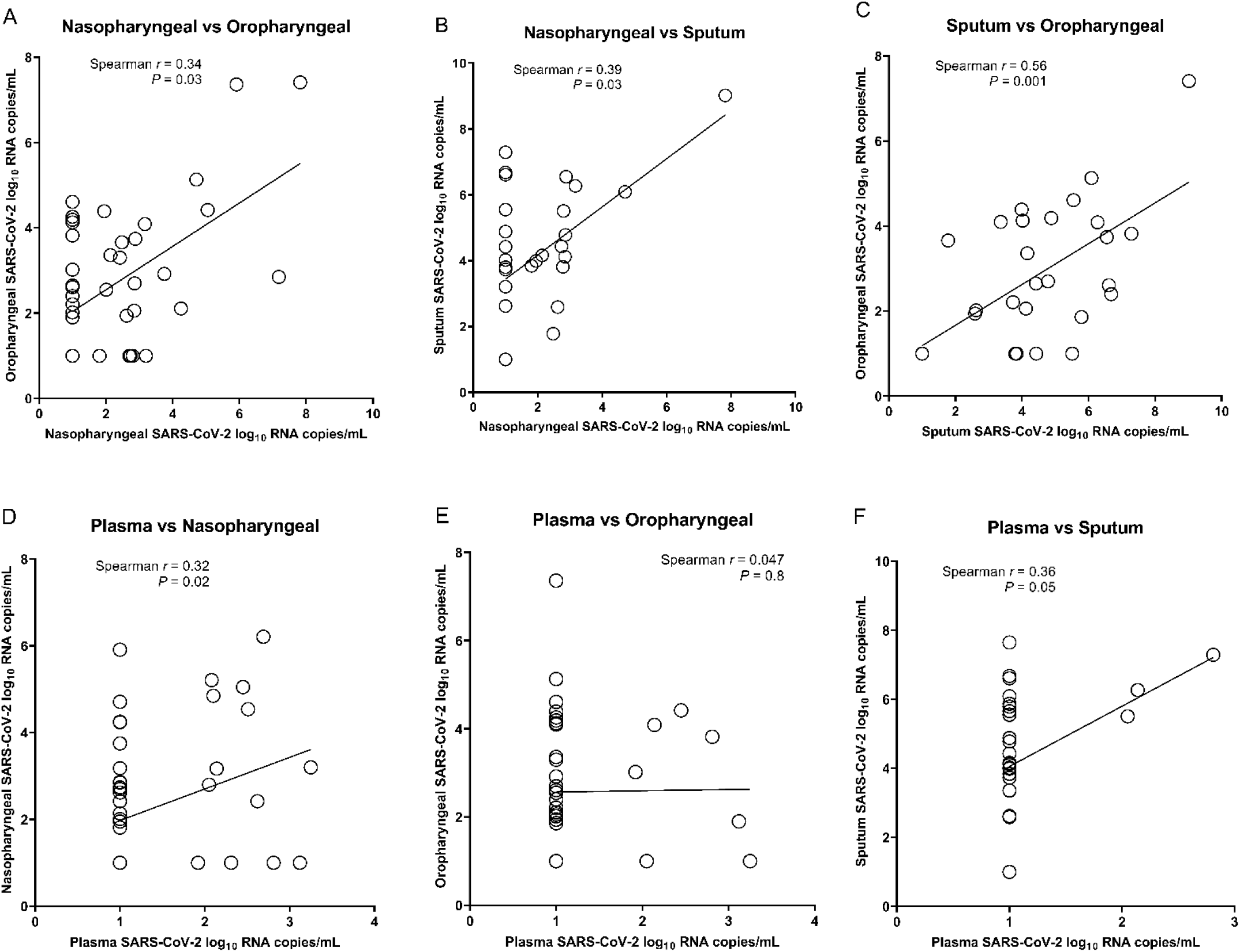
Correlation of respiratory tract and plasma viral loads. VL, viral load.

**Supplemental Figure 2.**
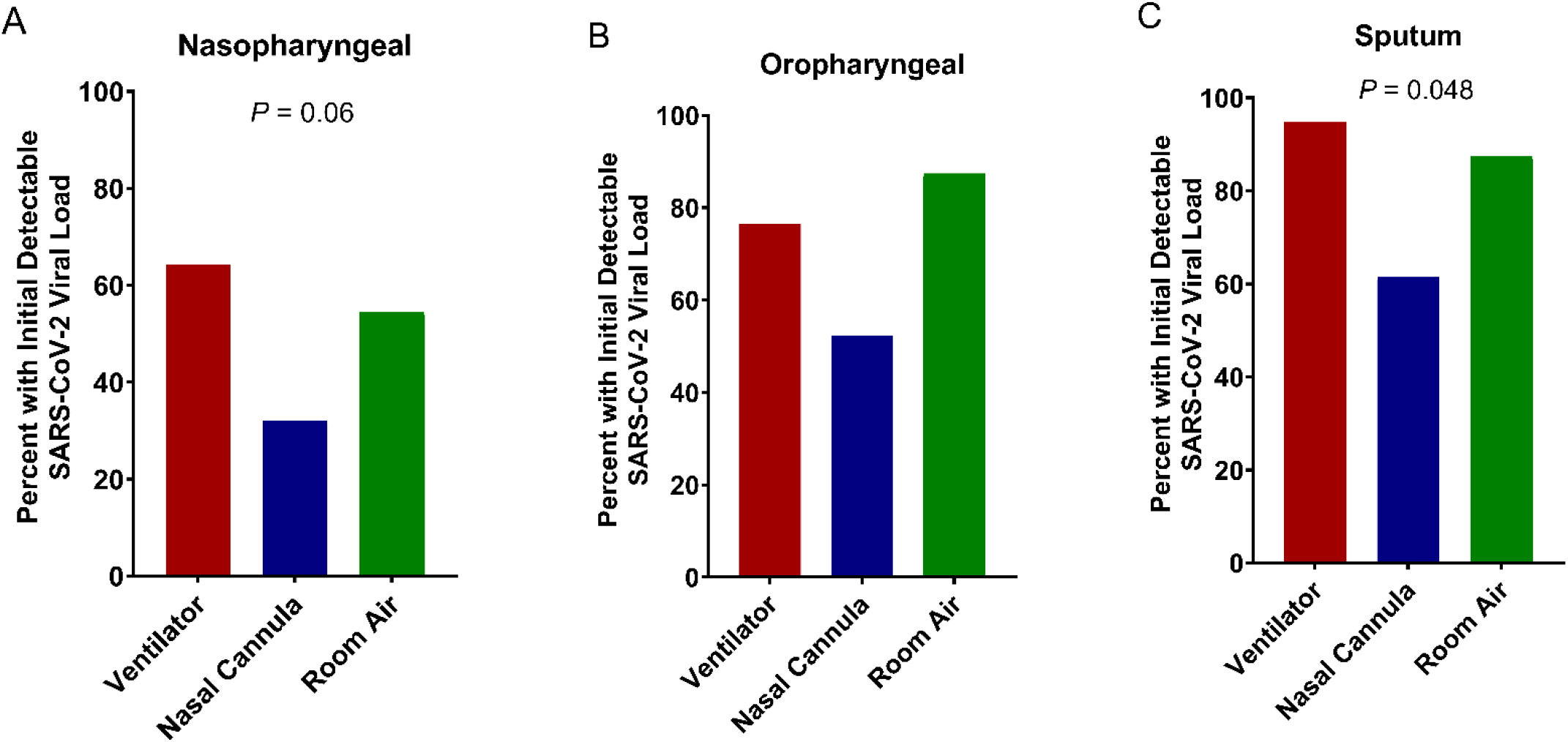
Detection of SARS-CoV-2 viral load from nasopharyngeal swabs (A), oropharyngeal swabs (B) and sputum (C), categorized by respiratory disease severity. P-values are from X^2^ analysis.

**Supplemental Table 1.** Logistic regression analysis of association of viral load with risk of death.

| **Specimen Type** | **Variable type<sup>1</sup>** | **Odds Ratio** | **P-value** |
| --- | --- | --- | --- |
| Plasma | Categorical | 5.5 | **0.02** |
| Nasopharyngeal | Categorical | 2.4 | 0.25 |
| Oropharyngeal | Categorical | 3.5 | 0.27 |
| Sputum<sup>2</sup> | Categorical | - | - |
| Plasma | Continuous | 2.4 | **0.04** |
| Nasopharyngeal | Continuous | 1.4 | 0.09 |
| Oropharyngeal | Continuous | 2.1 | **0.02** |
| Sputum | Continuous | 2.8 | **0.048** |

<sup>1</sup>Categorical refers to analysis of viral loads as detectable or not detectable
<sup>2</sup>There were no deaths in participants with undetectable sputum viral loads

